# Prevalence and associated factors of premarital sexual behavior among undergraduate youths of management colleges of Dhangadhi Sub-Metropolitan city, Kailali, Nepal

**DOI:** 10.1101/2024.05.23.24307834

**Authors:** Princy Bhatta, Min Bahadur Kunwar, Arun Kumar Joshi, Dharma Dev Bhatta, Ankit Acharya

## Abstract

**Background:** Although sexuality remains a taboo in Nepali society, the prevalence of premarital sexual activities is increasing. Youth who begin early premarital sexual activity are more likely to be engaged in unsafe sex. This study aims to determine the prevalence and associated factors of premarital sex among college-going youths of Dhangadhi Sub-Metropolitan City, Kailali (DSMC).

**Methodology:** This descriptive cross-sectional study was conducted in management colleges with Bachelor of Business Studies (BBS) programs in DSMC. The sample size for the study was 374. The probability proportionate to size sampling technique was adopted for sample estimation for the selected colleges. The data was entered, cleaned, and analyzed in SPSS26. Bivariate analysis was done using Pearson’s chi-square test was used to determine differences between groups. A simple logistic regression model was conducted to assess the association between several characteristics of the respondents and the outcome variable- premarital sex, followed by multiple logistic regression to evaluate the combined impacts of various predictors on premarital sex.

**Results:** The prevalence of premarital sex among undergraduate college youths was 35.3%, of which 52.3% were males. Among those undergraduate youths, more than half (51.5%) had experienced their first intercourse before 18 years of age. At least 1 out of 10 respondents reported that their intercourse led to pregnancy. In model I, in simple logistic regression, predictors such as age, gender, relationship status, dating experience (p-value < 0.001), communication with parents (p-value = 0.002), family type (p-value = 0.020), and family income (p-value = 0.002) were significantly associated with premarital sex. Furthermore, not living with parents, parent’s education level, friends with sex partners, friends having dating experience, and awareness that friends have sex were also among the factors significantly associated with premarital sex. In the multiple logistic regression, in Model II, relationship status (aOR, 44.47; 95% CI: 7.88, 251.08), dating experience (aOR, 24.00; 95% CI: 5.36, 107.58), discussion about sexual health with parents (aOR, 8.16; 95% CI: 2.25, 29.62), and knowing friends who had premarital sex (aOR, 15.82; 95% CI: 2.92, 85.61) were significantly associated with premarital sex after controlling for potential confounding predictors.

**Conclusion:** Sex education interventions within and beyond educational institutions are crucial to increase the level of awareness and protect the physical, mental, and social health of young unmarried partners in Nepal.

## Introduction

In 2018, the global youth population was 1.2 billion, with over 490 million young people in South Asia (1). In Nepal, young people aged 15-24 years constitute 19.71% of the total population (2). This young group of population demands high-quality sexual health care to address their specific sexual health needs and rights. Yet, poor understanding and awareness of sexuality and well-being complemented by limited access to sexual and reproductive health (SRH) services have been a major challenge for youths in low-and-middle-income countries (LMICs) to ensure a healthy transition to adulthood (3). Societies in LMICs across the world, where the majority of the youths reside, often associate immorality with young people’s sexuality and sexual practices before marriage. Because of these, young people are more likely to engage in risky sexual behaviors and may suffer physically, mentally, psycho-socially, and financially from its outcomes. Not only this, taboos related to sexuality are one of the major factors for the communication gap between youths and adults in those societies (3–5).

Premarital sex, also known as non-marital sex, youthful sex, adolescent sex, and young-adult sex, refers to sexual activity practiced before a couple is married (6). Advancements in science and technological innovations resulted in increased access to mobile phones with the internet, making it easy for young people to have social and romantic interactions (7, 8). Nepal Adolescents and Youth Survey 2010/11 reports that 12.98% of young people aged 15-24 years are involved in premarital sex (23.4% boys and 3.66% girls) (9). According to the Nepal Demographic and Health Survey 2022, 2% of never-married women and 25% of never-married men aged 15-24 had premarital sexual experience. The proportion of premarital sex has increased from less than 1% in 2006 to 2% in 2022 among never-married women, and from 17% in 2006 to 25% in 2016 and 2022(10). The recent demographic survey also highlights that 9% of women and 2% of men had their first sexual intercourse by age 15, i.e., before the legal age of marriage(10).

Sexual behavior including premarital sex is still a sensitive topic under discussion in Nepali society (9). Therefore, youths are not adequately informed about the right choices to prevent themselves and their partners from several negative health outcomes such as sexually transmitted infections (STIs), human immune virus and acquired immune deficiency syndrome (HIV and AIDS), teenage pregnancy, unsafe abortions, mental distress, and maternal deaths among many others (3, 11–13). Given this situation, there is a need to understand the factors that lead to premarital sexual behavior among young people to develop and implement interventions that are contextually appropriate and accepted. This study aims to assess factors affecting premarital sex among college-going youths of Dhangadhi Sub-Metropolitan City, Kailali (DSMC).

## Materials and methods

### Study design, respondents, and setting

This cross-sectional study was conducted among 18-24 years Bachelor in Business Studies (BBS) program students of Dhangadhi Sub-Metropolitan City (DSMC), Kailali from July 18, 2022, to July 26, 2022.

### Sampling frame, sample size, and sampling procedure

A multistage proportionate simple random sampling technique was used in this study. At first, the sampling frame was defined through a complete enumeration of all 4 colleges having BBS programs within DMPC. The sample size was determined using the sample size calculation method for proportions with finite population correction. The sample size from each college was obtained using the probability proportionate to size technique. The lottery method was used to select the academic year (1^st^ 2^nd^ or 3^rd^ or 4^th^) for data collection. For those academic years with multiple classrooms, a classroom was selected by using a lottery method.

#### Sample size calculation

The sample size for this study was 374, which was determined using the following formula with finite population correction:

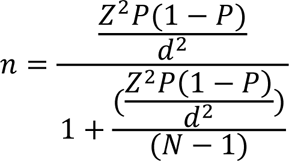

Where,

n = desired sample size

z = standard normal variable at 95% Confidence Level i.e. (1.96)

p = proportion in the target population estimated to have particular characteristics (according to the study on premarital sexual behavior in 2021, 38.1%college youths havehad premarital sex)

d = degree of accuracy required, set at 0.05 level

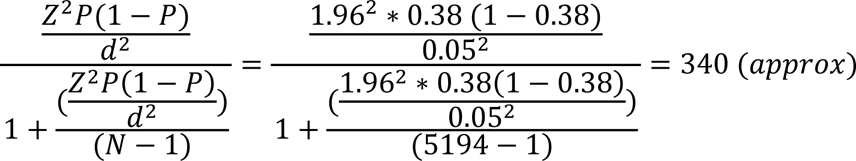

Assuming the non-response of 10%, the final sample size was 340 + 34 = 374.

### Data Collection Technique and Tools

The principal investigator of the study did an extensive literature review to develop a structured questionnaire. This was followed by incorporating experts’ opinions to ensure the completeness and quality of the tool. The questionnaire was then pretested among 10% of the total sample size at another college of Dhangadhi, Kailali where data collection was not planned. The pretested questionnaire was modified accordingly, distributed among all consenting youths (374), and collected immediately after completing the survey.

### Data Management and Analysis Plan

All questionnaire forms were gathered and checked for completeness and accuracy, and to ensure that they were ready for entry purposes. Data entry was done in SPSS V26, followed by data cleaning and data analysis. A backup of the original dataset was done and stored in a backup storage device and online storage system for future retrieval and use. Descriptive analysis was performed to describe the characteristics of the respondents. Bivariate analysis was done using Pearson’s chi-square test was used to determine differences between groups. A simple logistic regression model was conducted to assess the association between several characteristics of the respondents and the outcome variable-premarital sex. This was followed by multiple logistic regression to assess the combined impacts of multiple predictors on premarital sex. Model I for crude odds ratio and model II for adjusted odds ratios were reported, along with 95% Confidence Intervals (95% CIs). Ρ-value < 0.05 was considered to be statistically significant.

### Ethical Consideration

The ethical approval for this study was obtained from the Institution Review Committee (IRC) at Nobel College, Pokhara University. Before data collection, letters from the Department of Public Health at Nobel College were provided to all the colleges from where samples were collected. After the completion of data collection, a letter of data collection completion was obtained from each college. The objectives of the research, potential harm and benefit to the respondents were explained to all respondents along with the approximate time required for the interview, confidentiality, anonymity of data obtained, and the option to withdraw from the interview at any time if they felt uncomfortable. Written consent was taken from each respondent who agreed to participate in the study.

## Results

A total of 374 respondents participated in this study. The mean age and standard deviation of the respondents were 20.37 years and 1.164 years respectively. Almost three-fifths (59.4%) of respondents were male. The majority of the respondents (92.8%) followed Hinduism. More than one-third of the respondents represented Chhetri ethnicity, and nearly one-third of the respondents belonged to the Brahman community. The majority of the respondents (69.0%) were currently pursuing the second year of their undergraduate studies. Majority of the respondents (69.8%) were single. Out of 5, nearly 2 respondents (39.7%) received daily pocket money from their parents. Most of the respondents (96.0%) didn’t discuss sexual health with parents. More than half of the respondents (60.3%) never had a dating experience in their life.

Among all respondents, more than half (62.6%) belonged to the nuclear family. More than half of the respondent’s family income was between 20,000-30,000. The majority of the respondents (69.5%) lived with their parents. In regards to parents’ educational status, nearly one-third of mothers were not able to read and write (32.4%) while all fathers were able to. Nearly one-half of respondents’ fathers had a secondary or higher level of education (48.3%) as compared to only 14.2% of mothers. More than half (50.4%) of respondent’s father’s occupations were in agriculture and the majority (73.0%) of respondent’s mothers were engaged in household work. The majority (96.3%) of respondents’ parental marital status was married.

More than half (55.5 %) of the respondents do not consume any alcohol products. Among the drinkers, the majority (77.1%) of the respondents’ drink alcohol only during festivals. The majority of the respondents (75.1%) do not smoke tobacco products. Among the smokers, more than half (53.8%) of the respondents smoke only during festivals, followed by 22.6% of the respondents smoking frequently and 16.15 smoking regularly. The majority (94.7%) of the respondents do not consume drugs. More than half (56.7%) of the respondents watched porn movies. Among those who watched porn movies, 1 out of 10 respondents (11.8%) watched it every day. Almost two-thirds (63.6%) of the respondents perceived it was okay to have premarital sex. Table 1 shows the individual, family, and behavioral characteristics of the respondents.

**Table 1:**
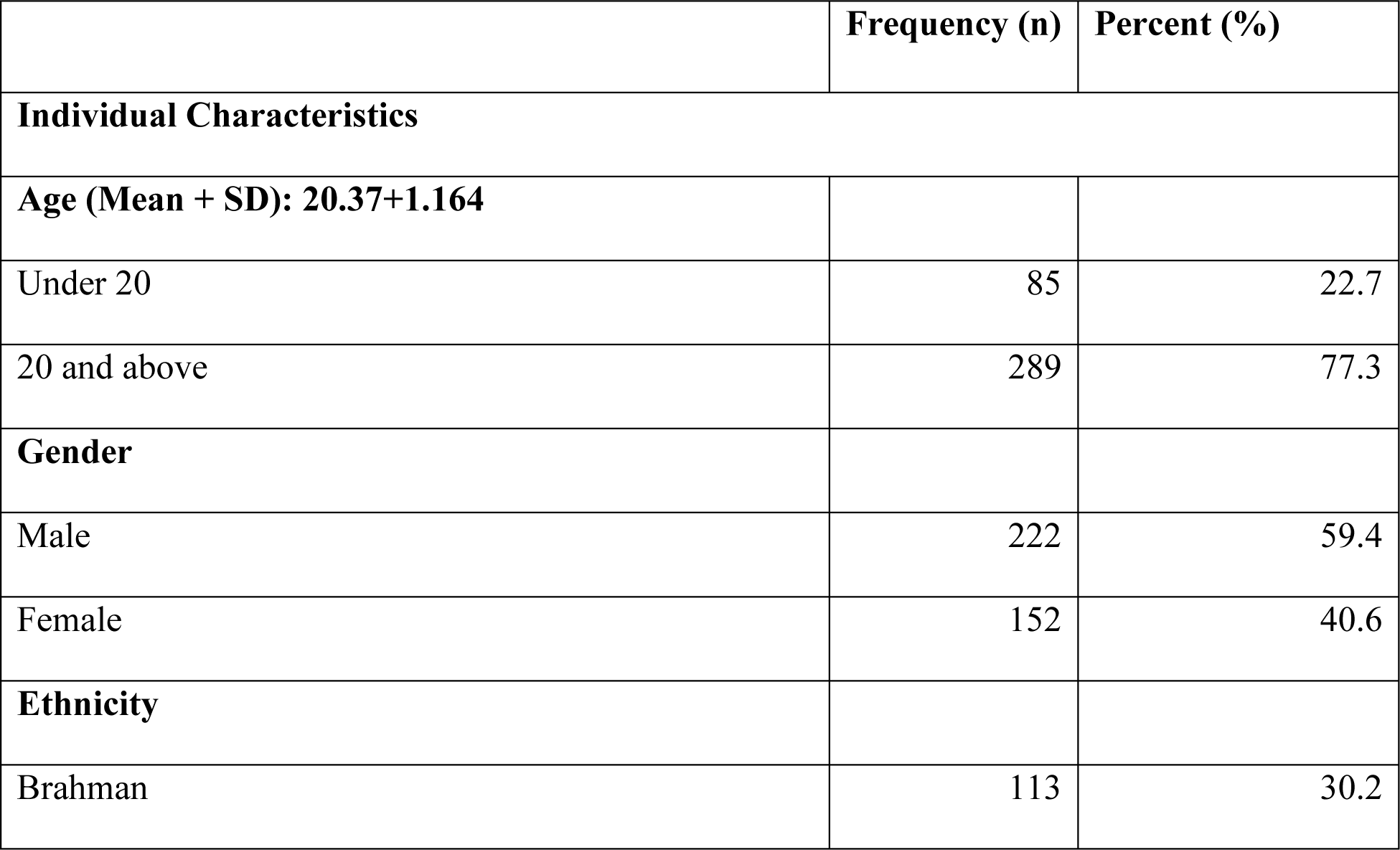
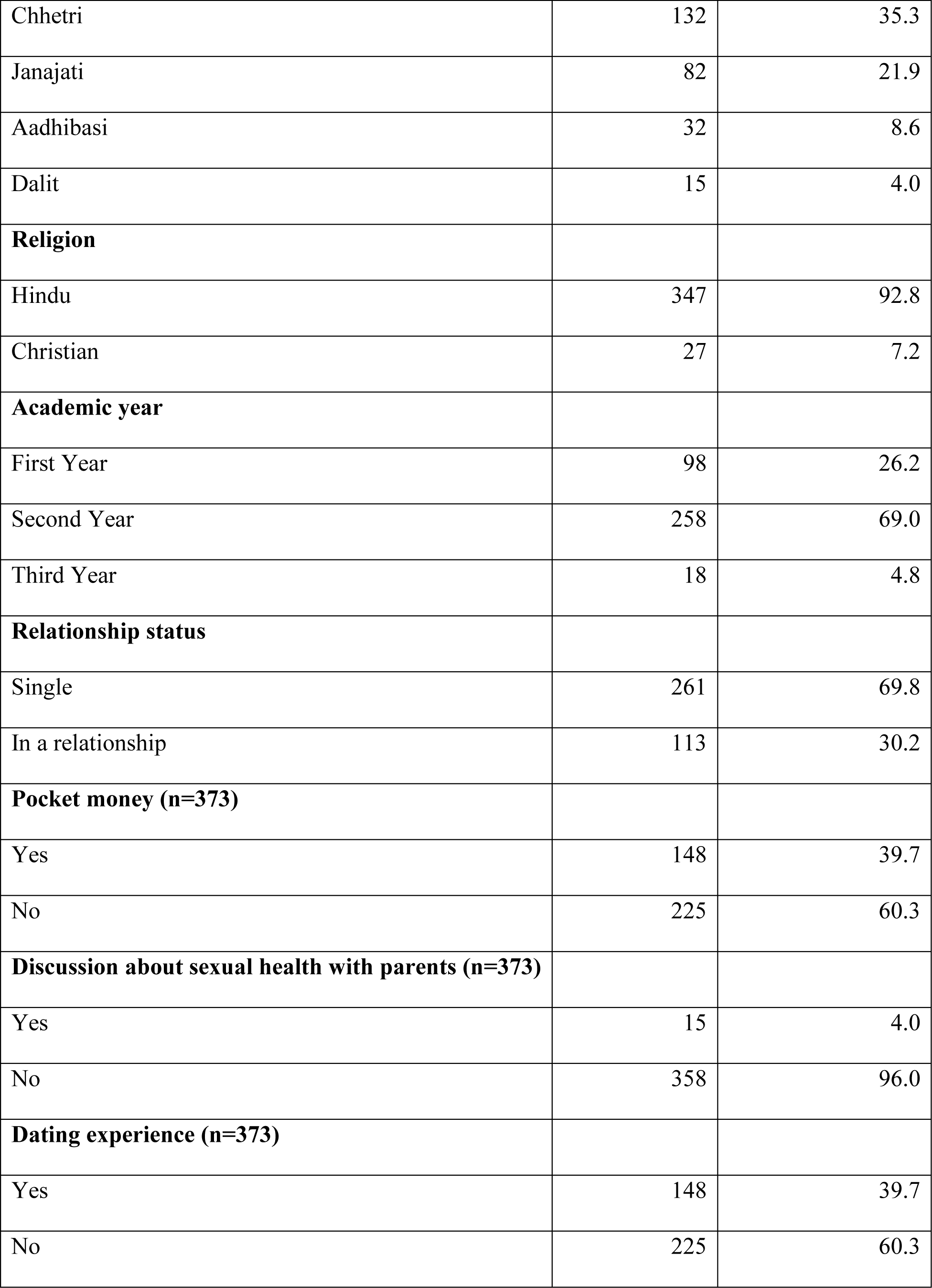
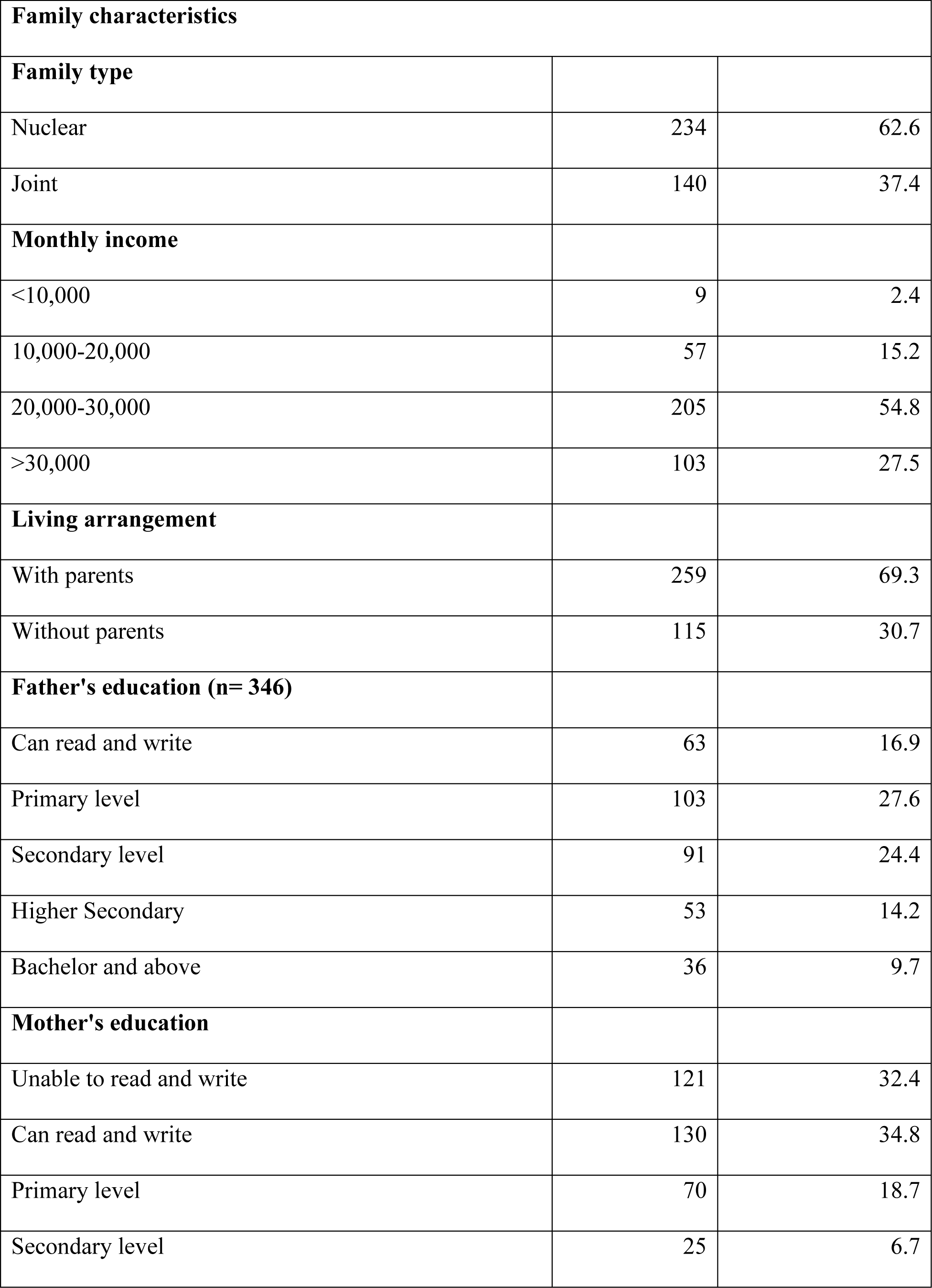
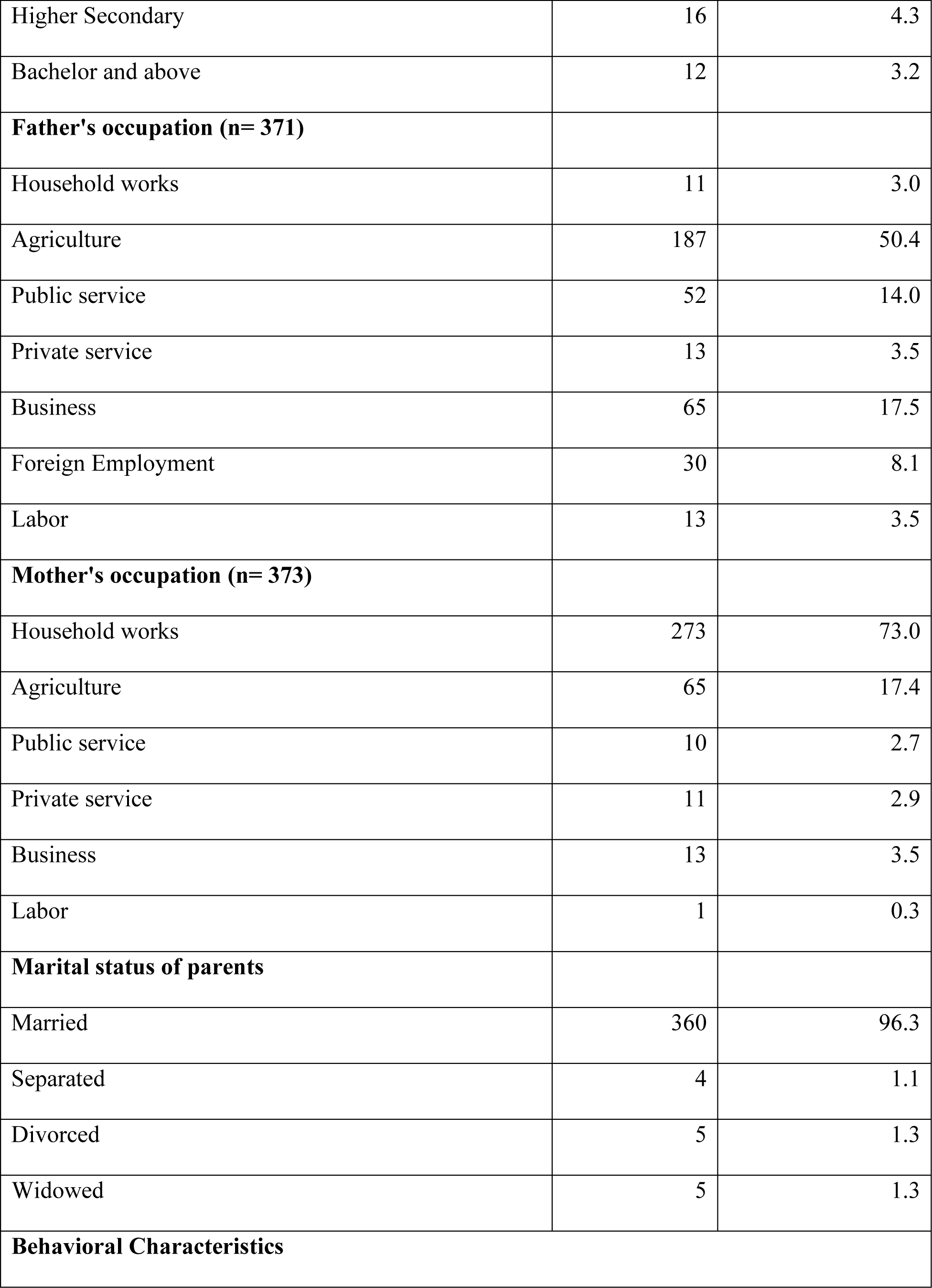
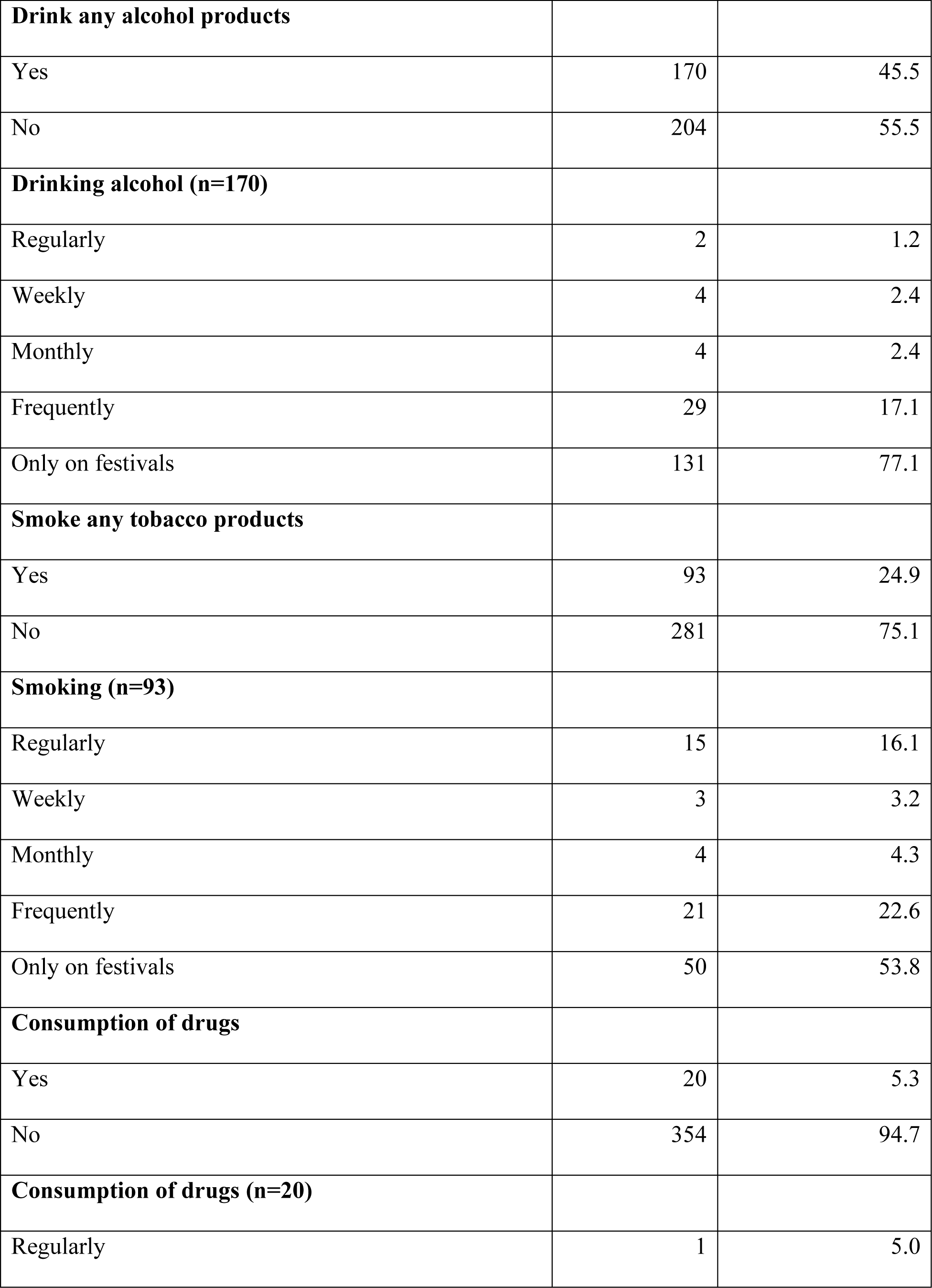
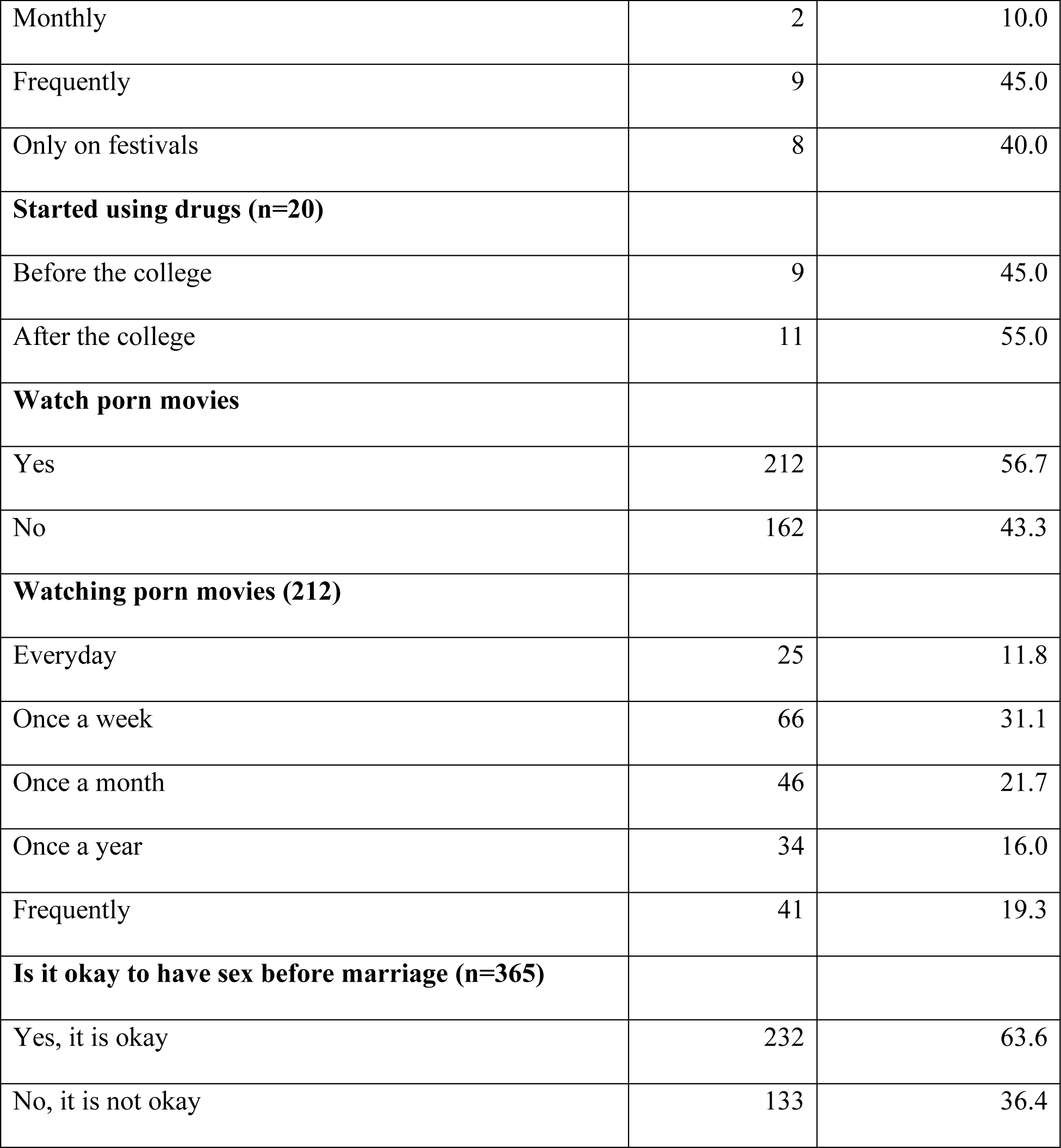
Individual, family, and behavioral characteristics of the respondents (n=374)

|  | Frequency (n) | Percent (%) |
| --- | --- | --- |
| **Individual Characteristics** |  |  |
| **Age (Mean + SD): 20.37+1.164** |  |  |
| Under 20 | 85 | 22.7 |
| 20 and above | 289 | 77.3 |
| **Gender** |  |  |
| Male | 222 | 59.4 |
| Female | 152 | 40.6 |
| **Ethnicity** |  |  |
| Brahman | 113 | 30.2 |
| Chhetri | 132 | 35.3 |
| Janajati | 82 | 21.9 |
| Aadhibasi | 32 | 8.6 |
| Dalit | 15 | 4.0 |
| **Religion** |  |  |
| Hindu | 347 | 92.8 |
| Christian | 27 | 7.2 |
| **Academic year** |  |  |
| First Year | 98 | 26.2 |
| Second Year | 258 | 69.0 |
| Third Year | 18 | 4.8 |
| **Relationship status** |  |  |
| Single | 261 | 69.8 |
| In a relationship | 113 | 30.2 |
| **Pocket money (n=373)** |  |  |
| Yes | 148 | 39.7 |
| No | 225 | 60.3 |
| **Discussion about sexual health with parents (n=373)** |  |  |
| Yes | 15 | 4.0 |
| No | 358 | 96.0 |
| **Dating experience (n=373)** |  |  |
| Yes | 148 | 39.7 |
| No | 225 | 60.3 |

| **Family characteristics** |  |  |
| --- | --- | --- |
| **Family type** |  |  |
| Nuclear | 234 | 62.6 |
| Joint | 140 | 37.4 |
| **Monthly income** |  |  |
| <10,000 | 9 | 2.4 |
| 10,000-20,000 | 57 | 15.2 |
| 20,000-30,000 | 205 | 54.8 |
| >30,000 | 103 | 27.5 |
| **Living arrangement** |  |  |
| With parents | 259 | 69.3 |
| Without parents | 115 | 30.7 |
| **Father's education (n= 346)** |  |  |
| Can read and write | 63 | 16.9 |
| Primary level | 103 | 27.6 |
| Secondary level | 91 | 24.4 |
| Higher Secondary | 53 | 14.2 |
| Bachelor and above | 36 | 9.7 |
| **Mother's education** |  |  |
| Unable to read and write | 121 | 32.4 |
| Can read and write | 130 | 34.8 |
| Primary level | 70 | 18.7 |
| Secondary level | 25 | 6.7 |
| Higher Secondary | 16 | 4.3 |
| Bachelor and above | 12 | 3.2 |
| **Father's occupation (n= 371)** |  |  |
| Household works | 11 | 3.0 |
| Agriculture | 187 | 50.4 |
| Public service | 52 | 14.0 |
| Private service | 13 | 3.5 |
| Business | 65 | 17.5 |
| Foreign Employment | 30 | 8.1 |
| Labor | 13 | 3.5 |
| **Mother's occupation (n= 373)** |  |  |
| Household works | 273 | 73.0 |
| Agriculture | 65 | 17.4 |
| Public service | 10 | 2.7 |
| Private service | 11 | 2.9 |
| Business | 13 | 3.5 |
| Labor | 1 | 0.3 |
| **Marital status of parents** |  |  |
| Married | 360 | 96.3 |
| Separated | 4 | 1.1 |
| Divorced | 5 | 1.3 |
| Widowed | 5 | 1.3 |
| **Behavioral Characteristics** |  |  |
| **Drink any alcohol products** |  |  |
| Yes | 170 | 45.5 |
| No | 204 | 55.5 |
| **Drinking alcohol (n=170)** |  |  |
| Regularly | 2 | 1.2 |
| Weekly | 4 | 2.4 |
| Monthly | 4 | 2.4 |
| Frequently | 29 | 17.1 |
| Only on festivals | 131 | 77.1 |
| **Smoke any tobacco products** |  |  |
| Yes | 93 | 24.9 |
| No | 281 | 75.1 |
| **Smoking (n=93)** |  |  |
| Regularly | 15 | 16.1 |
| Weekly | 3 | 3.2 |
| Monthly | 4 | 4.3 |
| Frequently | 21 | 22.6 |
| Only on festivals | 50 | 53.8 |
| **Consumption of drugs** |  |  |
| Yes | 20 | 5.3 |
| No | 354 | 94.7 |
| **Consumption of drugs (n=20)** |  |  |
| Regularly | 1 | 5.0 |
| Monthly | 2 | 10.0 |
| Frequently | 9 | 45.0 |
| Only on festivals | 8 | 40.0 |
| **Started using drugs (n=20)** |  |  |
| Before the college | 9 | 45.0 |
| After the college | 11 | 55.0 |
| **Watch porn movies** |  |  |
| Yes | 212 | 56.7 |
| No | 162 | 43.3 |
| **Watching porn movies (212)** |  |  |
| Everyday | 25 | 11.8 |
| Once a week | 66 | 31.1 |
| Once a month | 46 | 21.7 |
| Once a year | 34 | 16.0 |
| Frequently | 41 | 19.3 |
| **Is it okay to have sex before marriage (n=365)** |  |  |
| Yes, it is okay | 232 | 63.6 |
| No, it is not okay | 133 | 36.4 |

Among the respondents, more than one-third (35.5%) of the respondents had experienced premarital sexual experience. More than half (51.5%) had experienced premarital sex between the age of 14 and 18 years. More than one-half (50.8%) of the respondents had their first sex after college. About two-thirds (63.4%) of the respondent’s reason for first sexual intercourse was falling in love. More than 1 in 10 (12.1%) respondents reported that their involvement in sexual intercourse resulted in pregnancy. The majority (78.0%) of the respondents reported that they didn’t use condoms during their first sexual intercourse. Among non-users, more than half (52.5%) of the respondents reported that the trust factor played an important role in not using condoms during sexual intercourse. The majority (76.5%) of the respondents didn’t have sex with commercial sex workers while 23.5% had. Among those who had sex with commercial sex workers, the majority (74.2%) of the respondents used condoms. More than one-half (50.8%) of the respondents had only one sex partner, followed by more than one-third of the respondents (37.1%) having more than 3 or more partners.

Among the respondents who had a premarital sexual history, the majority (95.5%) of the respondents had premarital sex in the last 12 months (n=132). More than half (67.5%) of the respondents had one partner in the past 12 months. The majority (80.2%) of the respondents had sex with a boy/girlfriend in the past 12 months. More than three-fourths (77.0) of the respondents didn’t use condoms during sexual intercourse in the past 12 months. Nearly half (48.5%) of the respondents didn’t use condoms due to the reason of trusting their partner. The majority (84.8%) of the respondents didn’t have guilt of having premarital sex. Table 2 shows the premarital sexual history and the current sexual experience of the respondents.

**Table 2:** Premarital sexual history and current sexual experience of the respondents.

|  | **Frequency**<br>**(n)** | **Percent**<br>**(%)** |
| --- | --- | --- |
| **The premarital sexual history of the respondents (n=374)** |  |  |
| **Ever had premarital sex** |  |  |
| Yes | 132 | 35.3 |
| No | 242 | 64.7 |
| **Age at first intercourse** |  |  |
| 14-18 | 68 | 51.5 |
| 19-23 | 64 | 48.5 |
| **First sexual intercourse** |  |  |
| Before college | 65 | 49.2 |
| After college | 67 | 50.8 |
| **Reason for premarital sex** |  |  |
| Fall in love | 83 | 63.4 |
| Desire for sex | 47 | 35.9 |
| Peer pressure | 1 | 0.8 |
| **Ever become pregnant/partner pregnant (n=132)** |  |  |
| Yes | 16 | 12.1 |
| No | 116 | 87.9 |
| **Use a condom during first sexual intercourse** |  |  |
| Yes | 29 | 22.0 |
| No | 103 | 78.0 |
| **Reason for not using a condom (n=103)** |  |  |
| Ashamed to ask the partner | 7 | 6.8 |
| Ashamed to buy a condom | 2 | 1.9 |
| Trust partner | 53 | 52.5 |
| Partner objected | 8 | 7.8 |
| Used another contraceptive | 3 | 2.9 |
| Drunk | 4 | 3.9 |
| Decreased Satisfaction | 26 | 25.2 |
| **Sex with commercial sex workers** |  |  |
| Yes | 31 | 23.5 |
| No | 101 | 76.5 |
| **Use condoms with commercial sex workers (n=31)** |  |  |
| Yes | 23 | 74.2 |
| No | 8 | 25.8 |
| **Number of sex partners by now** |  |  |
| Only one | 67 | 50.8 |
| Two | 16 | 12.1 |
| Three and above | 49 | 37.1 |
| **Current premarital sexual experience of the respondents (n=132)** |  |  |
| **Have premarital sex in the past 12 months** |  |  |
| Yes | 126 | 95.5 |
| No | 6 | 4.5 |
| **Number of sex partners in the past 12 months (n=126)** |  |  |
| One | 85 | 67.5 |
| Two | 21 | 16.7 |
| Three | 20 | 15.9 |
| **With whom did you have sex in the past 12 months (n=126)** |  |  |
| Boy/Girl Friend | 101 | 80.2 |
| Commercial Sex worker | 11 | 8.7 |
| Teacher | 1 | 0.8 |
| Non-regular partner | 13 | 10.3 |
| **Use a condom during the last sexual intercourse (n=126)** |  |  |
| Yes | 29 | 23.0 |
| No | 97 | 77.0 |
| **Reason for not using a condom (n=97)** |  |  |
| Ashamed to Ask my partner | 5 | 5.2 |
| Trust partner | 47 | 48.5 |
| Partner objected | 5 | 1.3 |
| used other contraceptives | 1 | 0.3 |
| Drunk | 3 | 3.1 |
| Decreased satisfaction | 36 | 37.1 |
| **Have guilt of having premarital sex (n=132)** |  |  |
| Yes | 20 | 15.2 |
| No | 112 | 84.8 |

The majority (75.7%) of the respondents reported that their friends had boys/girlfriends. The majority (73.0%) of the respondents shared that they were aware of their friend’s dating experience. More than one-third (33.5%) of the respondents knew about the sexual intercourse of their friends. Among those who had sex, the majority (67.4%) of the respondents reported that their friends suggested they initiate their sexual intercourse. More than one-fifth (22.1%) of the respondents reported that their sexual behavior was influenced by girls’ clothing styles. Table 3 shows the peer influence on the respondents.

**Table 3:** Peer influence of the respondents (n=374)

| **Peer Influence** | **Frequency(n)** | **Percent (%)** |
| --- | --- | --- |
| **Friends have boy/girlfriend** |  |  |
| Yes | 283 | 75.7 |
| No | 91 | 24.3 |
| **Friends have dating experience** |  |  |
| Yes | 273 | 73.0 |
| No | 101 | 27.0 |
| **Know that your friends have experienced sexual intercourse (n=373)** |  |  |
| Yes | 125 | 33.5 |
| No | 248 | 66.5 |
| **Friends suggested you initiate your sexual intercourse (n=132)** |  |  |
| Yes | 89 | 67.4 |
| No | 43 | 32.6 |
| **Girls Wearing style influence your behavior (n=222)** |  |  |
| Yes | 49 | 22.1 |
| No | 173 | 77.9 |

The association between premarital sex and individual, family, behavioral, and peer influence characteristics was assessed using bivariate logistic regression, in model I. Age, gender, academic year, relationship status, dating experience, family type, living arrangement, discussion about sexual health with parents, drinking any alcohol products, smiling any tobacco products, consumption of drugs, watching porn, friends with boyfriends/girlfriends, and friends who had premarital sex were among the predictors that were significantly associated with premarital sex (p-value ≤ 0.05). In the multiple logistic regression, in Model II, relationship status, dating experience, discussion about sexual health with parents, and knowing friends who had premarital sex were significantly associated with premarital sex after controlling for potential confounding predictors, (p-value ≤ 0.05). In reference to those youths who were singly by relationship status, youths who were in a romantic relationship had 44.47 higher odds of having premarital sex (aOR, 44.47; 95% CI: 7.88, 251.08). In reference to those who had no dating experience, youths who had dating experience had 24 higher odds of having premarital sex (aOR, 24.00; 95% CI: 5.36, 107.58). In reference to those who lived with parents, those who lived without parents had 8.16 higher odds of having premarital sex (aOR, 8.16; 95% CI: 2.25, 29.62). Similarly, in reference to those who had no idea if their friends had had sex, those who knew their friends had had sex with their romantic partner had 15.82 higher odds of having premarital sex (aOR, 15.82; 95% CI: 2.92, 85.61). However, after the adjustments, associations were no longer statistically significant for substance abuse (drinking alcohol, smoking tobacco, consuming drugs) and watching pornographic movies. Similar was true for friends with boyfriend/girlfriend, and knowing friends who date their partners.

**Table 4:** Premarital sexual behavior and associated factors among undergraduate youth of management colleges of Dhangadhi Sub-Metropolitan city, Kailali, Nepal (n=374)

| Independent Variables | Dependent variable<br>(Premarital sex) |  | <sup>a</sup>Model I; cOR (95%<br>CI) | <sup>b</sup>Model II; aOR<br>(95% CI) |
| --- | --- | --- | --- | --- |
|  | Yes n (%) | No n (%) |  |  |
| AGE |  |  |  |  |
| Under 20 (85) | 15 (17.6) | 70 (82.4) | Reference category | Reference category |
| 20 and above<br>(289) | 117 (40.5) | 172 (59.5) | **3.17 (1.73, 5.81) \*** | 1.19 (0.16, 8. 60) |
| Gender |  |  |  |  |
| Male (222) | 116 (52.3) | 106 (47.7) | **9.30 (5.20, 16.63)\*** | 3.214 (0.40, 25.59) |
| Female (152) | 16 (10.5) | 136 (89.5) | Reference category | Reference category |
| Ethnicity |  |  |  |  |
| Brahman (113) | 33 (29.2) | 80 (70.8) | Reference category |  |
| Chhetri (132) | 45 (34.1) | 87 (65.9) | 1.25 (0.73, 2.16) |  |
| Janajati (82) | 32 (39.0) | 50 (61.0) | 1.55 (0.85, 2.83) |  |
| Aadhibasi (32) | 14 (43.8) | 18 (56.3) | 1.89 (0.84, 4.23) |  |
| Dalit (15) | 8 (53.3) | 7 (46.7) | 2.77 (0.93, 8.26) |  |
| Religion |  |  |  |  |
| Hindu (347) | 120(34.6) | 227(65.4) | Reference category |  |
| Christian (27) | 12(44.4) | 15(55.6) | 1.51 (0.69, 3.34) |  |
| Academic year |  |  |  |  |
| First year (98) | 23 (23.5) | 75 (76.5) | Reference category | Reference category |
| Second year<br>(258) | 96 (37.2) | 162 (62.8) | **1.93 (1.14, 3.29)\*** | 1.61 (0.33, 7.85) |
| Third year (18) | 13 (72.2) | 5 (27.8) | **8.48 (2.73, 26.31)\*** | 3.17 (0.07, 137.73) |
| Relationship status |  |  |  |  |
| Single (261) | 30 (11.5) | 231 (88.5) | Reference category | Reference category |
| In a relationship<br>(113) | 102 (90.3) | 11 (9.7) | **71.40 (34.44, 148.03)\*** | **44.47 (7.88, 251.08)\*** |
| Pocket money status |  |  |  |  |
| Yes (148) | 60 (40.5) | 88 (59.5) | 1.45 (0.94, 2.23) |  |
| No (225) | 72 (32.0) | 153 (68.0) | Reference category |  |
| Dating Experience |  |  |  |  |
| Yes (148) | 124 (83.8) | 24 (16.2) | **140.15 (61.11, 321.40)\*** | **24.00 (5.36, 107.58)\*** |
| No (225) | 8 (3.6) | 217 (96.4) | Reference category | Reference category |
| Family Type |  |  |  |  |
| Nuclear (234) | 93 (39.7) | 141 (60.3) | **1.71 (1.09, 2.69)\*** | 2,29 (0.61, 8.70) |
| Joint (140) | 39 (27.9) | 101 (72.1) | Reference category | Reference category |
| Living arrangement |  |  |  |  |
| With parents<br>(259) | 48 (18.5) | 211 (81.5) | Reference category | Reference category |
| Without parents<br>(115) | 84 (73.0) | 31 (27.0) | **11.91 (7.10, 19.99)\*** | **8.16 (2.25, 29.62)\*** |
| Discussion about sexual health with parents (n=373) |  |  |  |  |
| Yes (15) | 11 (73.3) | 4 (26.7) | **5.39 (1.68, 17.27)\*** | 3.31 (0.25, 44.50) |
| No (358) | 121 (33.8) | 237 (66.2) | Reference category | Reference category |
| Marital status of parents |  |  |  |  |
| Married (360) | 124 (34.4) | 236 (65.6) | Reference category |  |
| Separated/<br>Divorced/<br>Widowed (14) | 8 (57.1) | 6 (42.9) | 2.54 (0.86, 7.48) |  |
| Father's education (n=373) |  |  |  |  |
| Educated (90) | 33 (36.7) | 57 (63.3) | 1.08 (0.66, 1.76) |  |
| Uneducated<br>(283) | 99 (35.0) | 184 (66.7) | Reference category |  |
| Mother's Education |  |  |  |  |
| Educated (251) | 91 (36.3) | 160 (63.7) | 1.14 (0.72, 1.79) |  |
| Uneducated<br>(123) | 41 (33.3) | 82 (66.7) | Reference category |  |
| Family Income |  |  |  |  |
| <10,000 (9) | 2 (22.2) | 7 (77.8) | Reference category |  |
| 10,000-20,000<br>(57) | 19 (33.3) | 38 (66.7) | 1.75 (0.33, 9.25) |  |
| 20,000-30,000<br>(205) | 59 (28.8) | 146 (71.2) | 1.41 (0.29, 7.01) |  |
| More than<br>30,000 (103) | 52 (50.5) | 51 (49.5) | 3.57 (0.71, 18.00) |  |
| Drink any alcohol products |  |  |  |  |
| Yes (170) | 111<br>(65.3) | 59 (34.7) | **16.40 (9.50, 28.45)\*** | 1.67 (0.37, 7.47) |
| No (204) | 21 (10.3) | 183 (89.7) | Reference category | 1 |
| **Smoke any tobacco products** |  |  |  |  |
| Yes (93) | 70 (75.3) | 23 (24.7) | **10.75 (6.21, 18.61)\*** | 3.01 (0.63, 14.28) |
| No (281) | 62 (22.1) | 219 (77.9) | Reference category | Reference category |
| **Consumption of Drugs** |  |  |  |  |
| Yes (20) | 15 (75.0) | 5 (25.0) | **6.08 (2.16, 17.13)\*** | 7.44 (0.48, 115.11) |
| No (354) | 117<br>(33.1) | 237 (66.9) | Reference category | Reference category |
| **Watch porn movies** |  |  |  |  |
| Yes (212) | 128<br>(60.4) | 84 (39.6) | **60.19 (21.49, 168.56)\*** | 2.57 (0.32, 20.77) |
| No (162) | 4 (2.5) | 158 (97.5) | Reference category | Reference category |
| **Friends have boy/girlfriend** |  |  |  |  |
| Yes (283) | 124 (43.8) | 159 (56.2) | **8.09 (3.77, 17.35)\*** | 1.32 (0.05, 37.48) |
| No (91) | 8 (8.8) | 83 (91.2) | Reference category | Reference category |
| **Friends have dating experience** |  |  |  |  |
| Yes (273) | 123 (45.1) | 150 (54.9) | **8.382 (4.06,17.31)\*** | 0.26 (0.02, 4.54) |
| No (101) | 9 (8.9) | 92 (91.9) | Reference category | Reference category |
| **Know friends who had premarital sex (n=373)** |  |  |  |  |
| Yes (125) | 91 (72.8) | 34 (27.2) | **13.51 (8.06, 22.67)\*** | **15.82 (2.92, 85.61)\*** |
| No (248) | 41 (16.5) | 207 (83.5) | Reference category | Reference category |

\*p-value < 0.05 at 95% Confidence Level
<sup>a</sup>Model I, unadjusted odds ratio; <sup>b</sup>Model II, unadjusted odds ratio

## Discussion

This study found that the prevalence of premarital sex among undergraduate college youths in DMPC was 35.3%. Studies conducted in other parts of Nepal have reported a lower prevalence of premarital sex: Kathmandu 13.5% (14), Pokhara 24.6% (15), Jhapa 25% (16) and Web-based survey 21.2% (17). Among those who experienced premarital sex, one in two youths experienced it after they were enrolled in college and before the age of 18 years. This study also found that almost 4 of 5 respondents did not use condoms during their first sexual intercourse, and 1 of 10 sexual encounters led to pregnancy. Although adolescents report trust factor to be a cause for not using contraceptives during sex, the underlying reasons are logistic barriers such as lack of access to free condoms, financial dependence, and shyness to buy condoms from a health facility or pharmacy (18). This study found that almost 1 out of 4 youths had sex with commercial sex workers, and among those youths almost 3 out of 4 used condoms. This is possibly due to increased awareness of HIV and AIDS transmission among youths and to avoid the risk of transmission of STIs including HIV and AIDS (19).

This study found that nearly half of the youths had multiple sex partners with more than one-third of youths having more than 2 sex partners. Similar findings have been presented by Paudel and colleagues in their web-based study in Nepal (17) and by Upreti and Acharya (14) in Kathmandu. In this study, almost two-thirds of the respondents perceived that it was okay to have premarital sex, and among those who ever had premarital sex, the majority didn’t have any guilt about having premarital sex. This presents the changing dynamics of Nepali society on sex and sexuality as youths nowadays do not hesitate to make boyfriends/girlfriends, stay in romantic relationships, and enjoy sex life before marriage without regrets. Although Nepali society has accepted these kinds of behavior with increasing awareness of sexual freedom and rights, such practices may lead to an increased risk of unwanted pregnancy, maternal death, transmission of STIs, mental distress, violence, and suicide. Therefore, increased investment, awareness, and partnership for expanding sex education programs and interventions is crucial to make sure that youths make informed and meaningful decisions in their lives.

Peer influence is one of the major determinants for initiation of sexual activities among adolescents and youths. In this study, more than two-thirds of the respondents shared that they were aware of their friends’ dating experiences, and more than one-third knew about their friends’ sexual relationships. Not only this, among those who already had premarital sexual intercourse, more than two-thirds reported that it was their friend/s who suggested they initiate sex. This depicts that, friends, who are an important source of sex education in Nepali society, can influence the intention to initiation of sexual activities since discussing sex and sexuality with parents and other social networks is still not common (only 4% of youths discussed sexual health with parents) (20, 21). Predictors such as age, gender, relationship status, dating experience, communication with parents, family type, not living with parents, parent’s education level, friends with boyfriend/girlfriend, friends having dating experience, and awareness that friends have sex were also among the factors significantly associated with premarital sexual behavior. Several studies report that indulgence in substance abuse increases the likelihood of engaging in premarital sex (21, 22). Although watching pornographic content is illegal in Nepal following the government’s porn ban to reduce the cases of sexual violence including rape since September 2018 (23), more than one-half of youths still watch porn videos on the internet.

Being a retrospective study, recall bias might have occurred while collecting responses from the respondents. Socially desirable responding (SDR) cannot be controlled for sensitive research where private data is asked. Some important variables such as “having a personal laptop or mobile phone”, “media exposure”, and “access to SRHR services” were missed in the questionnaire, but could potentially contribute to premarital sexual behavior.

## Conclusion

This study revealed a high prevalence of premarital sex among college-going youths in DMPC, one of the vibrant cities of Sudurpaschim province in Nepal. This finding implies that there is a higher risk of STIs, HIV and AIDS, unwanted pregnancy, teenage pregnancy, abortion, violence, and mental health distress among the youths in Nepal. Discussing healthy relationships, and safer sex, and promoting sex education awareness programs within and beyond educational institutions is crucial to increase the level of awareness of young unmarried partners towards sexuality and well-being. Effective implementation of curriculum-based comprehensive sexuality education, and promoting peer-to-peer education as a method in sexual and reproductive health and rights promotion and risk communication among adolescents and youths must be encouraged by educational institutions or school-based clubs in Nepal. Incorporating creative and multi-channel approaches such as social and behavior change communication (SBCC) can influence positive behavior among youths and promote healthy indulgence in sexual activities. Investing in adolescents and youths, and improving access to sexual and reproductive health and rights is critical not only to prepare a healthy and capable youth force for the future but also to achieve the targets of sustainable development goals (SDGs).

## Data Availability

We have attached the data used in the paper as Supporting Information.

## Acknowledgements

We extend our profound gratitude to all faculties of Nobel College, Pokhara University and the research division of Sustainable Public Health Pvt. Ltd. for providing valuable guidance and insights in this study.

## References

1. Nations U. World population prospects 2019. Vol (ST/ESA/SE A/424) Department of Economic and Social Affairs: Population Division. 2019.

2. National Population and Housing Census 2021 (National Report). Ramshahpath,Thapathali, Kathmandu, Nepal; 2023.

3. Alam P, Lin L, Thakkar N, Thaker A, Marston C. Socio-sexual norms and young people’s sexual health in urban Bangladesh, India, Nepal and Pakistan: A qualitative scoping review. PLOS Glob Public Health. 2024;4(2):e0002179.

4. Van Reeuwijk M, Nahar P. The importance of a positive approach to sexuality in sexual health programmes for unmarried adolescents in Bangladesh. Reproductive health matters. 2013;21(41):69–77.

5. Nahar Q, Tuñón C, Houvras I, Gazi R, Reza M, Huq NL. Reproductive health needs of adolescents in Bangladesh: A study report: International Centre for Diarrhoeal Diseases Research Bangladesh: Dhaka; 1999.

6. Cavendish M. Sex and society. London: Marshall Cavendish Corporation. 2010:577.

7. McCormack M. The role of smartphones and technology in sexual and romantic lives. 2015.

8. Regmi PR, Simkhada P, van Teijlingen ER. There are too many naked pictures found in papers and on the net: Factors encouraging premarital sex among young people of Nepal. Health Science Journal. 2010;4(3):169–81.

9. Kathmandu N. Nepal Adolescents and Youth Survey. 2012.

10. Ministry of Health and Population [Nepal], New ERA, and ICF. 2023. Nepal Demographic and Health Survey 2022. Kathmandu, Nepal: Ministry of Health and Population [Nepal].

11. Adhikari N, Adhikari S. Attitude towards Premarital Sex among Higher Secondary Students in Pokhara Sub-Metropolitan City. J Community Med Health Educ 7: 564. doi: 10.4172/2161-0711.1000564 Page 2 of 6 J Community Med Health Educ, an open access journal ISSN: 2161-0711 Volume 7• Issue 5• 1000564. were in early adolescent stage ie age group of. 2017:14-6.

12. Shrestha RB. Premarital sexual behaviour and its impact on health among adolescents. Journal of Health Promotion. 2019;7(1):43–52.

13. Bhandari SD. Knowledge on Premarital Sex and its Consequences Among Adolescents at a Higher Secondary School. Medical Journal of Shree Birendra Hospital. 2014;13(1):37–9.

14. Upreti YR, Acharya D. Premarital Sexual Behaviours among Secondary School Adolescents: A Cross-sectional Study in Kathmandu. Journal of Health Promotion. 2020;8:39–52.

15. Adhikari N, Adhikari S, Sulemane NI. Premarital sexual behaviour among higher secondary students in Pokhara Sub-Metropolitan City Nepal. Sexual health. 2018;15(5):403–7.

16. Bhatta DN, Koirala A, Jha N. Adolescent students’ attitude towards premarital sex and unwanted pregnancy. Health Renaissance. 2013;11(2):145–9.

17. Paudel A, Neupane A, Khadka S, Adhikari L, Paudel S, Kaphle M. Factors affecting Premarital Sex among Nepalese Undergraduates. Journal of Public Health and Development. 2023;21(2):152–67.

18. Regmi PR, van Teijlingen E, Simkhada P, Acharya DR. Barriers to sexual health services for young people in Nepal. Journal of health, population, and nutrition. 2010;28(6):619.

19. Mahat G, Scoloveno MA. HIV/AIDS knowledge, attitudes and beliefs among Nepalese adolescents. Journal of advanced nursing. 2006;53(5):583–90.

20. Regmi P, van Teijlingen E, Silwal R, Dhital R. Role of social media for sexual communication and sexual behaviors: A focus group study among young people in Nepal. Journal of Health Promotion. 2022;10(1):153–66.

21. Dahal M, Subedi RK, Khanal S, Adhikari A, Sigdel M, Baral K, et al. Prevalence and Possible Risk Factor of Premarital Sexual Behaviour among Nepalese Adolescents. 2020.

22. Pahari S, Adhikari C. Premarital sexual behaviours of college youth of Tanahun district, Nepal. Journal of Health and Allied Sciences. 2021;11(1):30–7.

23. Karmacharya A. Ban your fears. My Republica. 2019.

